# The use and outcomes of non-pharmacological analgesia in the adult emergency department

**DOI:** 10.1101/2023.09.24.23296042

**Authors:** Aisha Amzaidy M Assiry, Nathan J Brown, Sarah Hazelwood, Anna-Lisa Lyrstedt, Rajeev Jarugula, Lee Jones, Kevin Chu, James A Hughes

## Abstract

**Background:** The treatment of pain in the emergency department (ED) has historically relied on pharmacological analgesia. However, little is known about the concurrent use of non-pharmacological analgesia.

**Aims/Objectives:** This research explores the self-reported use and outcomes of non-pharmacological analgesia in adult ED patients with moderate to severe pain.

**Methods:** This is a cross-sectional study in an adult ED of self-reported use of non-pharmacological pain management via a patient-reported outcome measure. The results are presented with descriptive statistics and multivariable modelling.

**Results:** 45.8% (n=296) of all respondents used non-pharmacological interventions. The most used non-pharmacological interventions are hot packs (34.9%, n=103), distraction (22.3%, n=66), and cold packs (12.9%, n=38). In multivariate modelling, females, patients who did not receive pre-hospital analgesia, and daily average access block time all affected the use of non-pharmacological analgesia. Within a multivariable model, non-pharmacological analgesia increased the amount of pain relief achieved.

**Conclusion:** Non-pharmacological analgesia use is affected by gender, treatment before the ED and ED workload. Nevertheless, clear benefits to the use of non-pharmacological analgesia were seen. Further work must be undertaken to encourage providers’ use of this modality and capture any additional benefits to the patient.

## Introduction

In Australia, pain is the most common presenting symptom in the emergency department (ED), with over 55% of patients presenting with pain (1). Inadequate pain management is common in the ED (2). To improve the quality and effectiveness of pain management in the ED, we need a better understanding of the utilisation and outcomes of current treatments. With increasing scrutiny of pharmacological analgesia, especially opiates (3), non-pharmacological analgesia represents an alternative or adjunctive therapy for many patients. However, little is known about the current prevalence of use, and effectiveness, of non-pharmacological analgesia in the ED.

The use of pharmacological analgesia is associated with increased risks of addiction and tolerance (4, 5). It has been suggested that multi-modal approaches that include non-pharmacological interventions are safe alternatives that can reduce over-reliance on pharmacological analgesia and its associated adverse effects (5-7). The numerous non-pharmacological options for pain management include, amongst other things, the application of heat or cold, activities that distract patients, meditation and massage. Evaluating the effectiveness of pain management in the ED has been difficult due to a lack of appropriate tools. The recent adaptation (8) and validation (9) of the Revised American Pain Society – Patient Outcome Questionnaire in the ED setting (APS-POQ-RED) provides the means to assess the usage and effectiveness of pain relief in the ED including non-pharmacological analgesia.

The primary aim, in this study of adult ED patients, was to determine the prevalence of use and characteristics of non-pharmacological analgesia for the treatment of pain in the ED. The secondary aim was to determine the effects of non-pharmacological analgesia on patient reported pain outcomes.

## Methods

### Research Design

This cross-sectional study uses data that was collected in a previous study, designed to psycometrically validate a new patient-reported outcome measure for acute pain in the ED (9). It is reported in line with the ‘Strengthening the reporting of observational studies in epidemiology’ (STROBE) checklist (10).

### Specific Objectives

The specific objectives, in this study of adult ED patients with moderate to severe pain, were:

1. To determine the use of non-pharmacological analgesia for the treatment pain
2. To compare the demographics, clinical characteristics and outcomes between patients who received pharmacological and non-pharmacological analgesia.
3. To determine the effect of non-pharmacologcial analgesia on pain intensity score

### Participants

Patients were eligible for inclusion if they were aged 18 years or older, had a pain intensity score ≥ 4/10 on arrival in the ED, and had the cognitive capacity for communication, written consent, and response to research questions. Patients were excluded if they had been transferred from another hospital, did not speak English, required resuscitation, or had a cognitive impairment through organic process or intoxication.

### Recruitment and Consent

The study was conducted in two large inner-city EDs within Australia’s largest health service between September 2021 and January 2022. A research assistant identified patients with acute pain, >3/10 in severity, from the electronic information system on Monday to Friday between 0700 and 1600. Potential participants were invited to partake after receiving ED care and before discharge or admission to an inpatient ward. Written informed consent to participate was obtained. Ethical approval was granted by the hospital’s Human Research Ethics Committee to conduct the study (LNR/2019/QRBW/55143).

### Instruments

This study used the ED adaptation of the Revised American Pain Society – Patient Outcome Questionnaire (APS-POQ-RED) to collect information on the types and outcomes of pain management in the ED (8, 9). The APS-POQ-RED also collects information about pain intensity, such as most intense pain experienced in the ED, least pain experienced, and the amount of relief received.

### Collected Data

In addition to the APS-POQ-RED, data about demographics (age, gender, socioeconomic status, comorbidities and country of birth), pharmacological analgesia provided, triage category and arrival and departure times, were also collected. Information about ED workload was collected because of its known influence on pain care (11). Variables collected were guided by previous applications of Symptom Management Theory (SMT) to pain care in the ED (11, 12).

### Outcome Measure

The primary outcome measure was whether a non-pharmacological intervention was used to treat an ED patient who was in pain. The secondary outcome measure was the level of pain relief achieved using all forms of analgesia.

### Sample Size

The sample was of sufficient size to determine a prevalence of at least 50% (+/-5%) with a confidence level of 95%. It was also sufficiently powered to conduct the multivariable regressions, providing at least 10 datapoints per independent variable (13).

### Statistical Analysis

The data were analysed using SPSS Version 27 (IBM Corp). Continuous variables were summarised using means and standard deviation and categorical variables using frequencies and percentages. Each patient was stratified into one of four groups according to the types and combinations of analgesia they received in the ED: ‘PA only’ = patients who received pharmacological analgesia only; ‘PA+NPA’ = patients who received both pharmacological and non-pharmacological analgesia; ‘NPA only’ = patients who received non-pharmacological analgesia only; ‘No Analgesia’ = patients who received neither pharmacological nor non-pharmacological analgesia. Between-group differences for continuous variables were determined using t-tests and analysis of variance (ANOVA) (14, 15), with the Bonferroni correction for multiple comparisons (16). Between-group differences for categorical variables were determined using the Chi-squared test of association. Binary logistic regression was used to determine the independent predictors of non-pharmacological analgesia use in the ED. Multivariable linear regression was used to determine the independent predictors of the change in pain intensity. Each regression model used a backwards stepwise method in which all candidate predictor variables with a p ≤ 0.25 were entered into the model, then, one-by-one the least significant variables were removed until each remaining predictor variable was significant to p < 0.05 (17).

## Results

Table 1 shows the demographic, presentation and clinical characteristics of participants stratified by treatment group. Of the 653 patients recruited, one withdrew, five were excluded due to only having mild pain (< 4/10), and one was excluded for being under 18 years at the time of recruitment. Thus, 646 patients were included in the analysis. Mean (SD) age was 48.3 (19.2) years, 55.6% were females. Classification by type of analgesia resulted in 322 (49.8%) participants in the PA only group, 270 (41.8%) in the PA+NPA group, 26 (4.0%) in the NPA only group and 28 (4.3%) in the No Analgesia group. Overall, 296 (45.8%) received non-pharmacological analgesia.

**Table 1:** Demographic, presentation and clinical characteristics of 646 adult ED patients with moderate to severe pain, stratified by treatment group.

|  | Units | Total | Treatment Group |  |  |  | p |
| --- | --- | --- | --- | --- | --- | --- | --- |
|  |  |  | PA only | PA+NPA | NPA only | No Analgesia |  |
| Number of patients | n (%) | 646 | 322 (49.8) | 270 (41.8) | 26 (4.0) | 28 (4.3) |  |
| Age | Mean years (SD) | 48.3 (19.2) | 48.8 (18.7) | 46.6 (19.0) | 53.5 (22.6) | 54.1 (22.0) | 0.080 |
| Males | n (%) | 283 (43.8) | 163 (50.6) | 96 (35.6) | 9 (34.6) | 15 (53.6) | 0.002 |
| Females | n (%) | 359 (55.6) | 158 (49.1) | 172 (63.7) | 17 (65.4) | 12 (42.9) |  |
| Born in Australia | n (%) | 474 (73.4) | 240 (74.5) | 192 (71.1) | 19 (73.1) | 23 (82.1) | 0.566 |
| Born elsewhere | n (%) | 172 (26.6) | 82 (25.5) | 78 (28.9) | 7 (26.9) | 5 (17.9) |  |
| Non-Indigenous | n (%) | 623 (96.4) | 313 (97.2) | 260 (96.3) | 24 (92.3) | 26 (92.9) | 0.407 |
| Indigenous Australian | n (%) | 23 (3.6) | 9 (2.8) | 10 (3.7) | 2 (7.7) | 2 (7.1) |  |
| ISRAD | Mean (SD) | 1045 (51.9) | 1048 (51.7) | 1041 (52.5) | 1033 (50.1) | 1055 (47.9) | 0.152 |
| CCI, age-adjusted | Mean (SD) | 2.0 (2.8) | 2.2 (2.9) | 1.7 (2.7) | 2.5 (2.3) | 2.9 (2.7) | 0.053 |
| New health problem | n (%) | 367 (56.8) | 189 (58.7) | 145 (53.7) | 13 (50.0) | 20 (71.4) | 0.502 |
| Accident or injury | n (%) | 135 (20.9) | 63 (19.6) | 63 (23.3) | 5 (19.2) | 4 (14.3) |  |
| Ongoing health condition | n (%) | 144 (22.3) | 70 (21.7) | 62 (23.0) | 8 (30.8) | 4 (14.3) |  |
| Pain intensity on arrival to ED<sup>\$</sup> | Mean score (SD) | 7.3 (2.0) | 7.1 (2.0) | 7.5 (1.9) | 6.6 (1.9) | 6.0 (2.2) | <0.001 |
| Highest pain intensity in ED<sup>\$</sup> | Mean score (SD) | 7.8 (1.9) | 7.9 (1.9) | 7.9 (1.8) | 7.0 (2.0) | 6.4 (2.0) | <0.001 |
| Lowest pain intensity in ED<sup>\$</sup> | Mean score (SD) | 2.3 (2.3) | 2.4 (2.3) | 2.3 (2.3) | 1.8 (2.9) | 1.1 (1.9) | 0.036 |
| % time with severe pain in ED | Mean % (SD) | 30.7 (32.9) | 34.3 (33.8) | 29.9 (32.3) | 14.6 (25.0) | 11.4 (21.0) | <0.001 |
| % pain relief in ED | Mean % (SD) | 69.7 (31.1) | 67.2 (31.9) | 70.4 (30.1) | 78.1 (30.5) | 82.9 (28.1) | 0.028 |

IRSAD = Index of relative socioeconomic advantage and disadvantage, CCI = Charlson Comorbidity Index, ED = Emergency Department, PA only = patients who received pharmacological analgesia only, PA+NPA = patients who received both pharmacological and non-pharmacological analgesia, NPA only = patients who received non-pharmacological analgesia only, No Analgesia = patients who received neither pharmacological nor non-pharmacological analgesia. \$ = pain intensity scoring out of 10. Additional variables used in multivariable modelling are described in Supplementary Table 1.

### Demographics

Mean age, country of birth, indigenous status, mean index of socioeconomic advantage and disadvantage (IRSAD) score, and mean Charlson comorbidity score (CCI) were similar between the four groups. Gender distribution differed across the four groups, with females having a greater odd of receiving pharmacological analgesia and non-pharmacological analgesia (PA+NPA) rather than pharmacological analgesia only (PA) (OR 1.848, 95%CI 1.326 – 2.580, p<0.001).

### Pain

The majority (90.6%, n=585) of the patients had pain intensity recorded on arrival. Mean pain scores on arrival varied between the four treatment groups. It was highest in the PA-NPA group and lowest in the NA group. The mean pain score for the worst pain experienced in the ED was slightly higher than for arrival pain. The difference in mean pain scores between arrival and worst pain experienced varied between treatment groups. It was higher in the PA+NPA groups and lower in the NPA+NA groups and were not significantly different from each other. There was a significant difference between those who received only pharmacological analgesia and those who received no analagesia.

There was a significant difference across the four groups in terms of the percentage of time in severe pain. The PA only and PA+NPA groups had similar percentages of time in severe pain (34.3% and 29.9%) and were not significantly different. The NPA only and No Analgesia groups had lower percentages of time in severe pain and were not significantly different from each other. The PA only group differed from the NPA only and No Analgesia groups.

Patients were asked to report on the amount of relief they received from their analgesia. There was a significant difference across the four groups. The PA only group and the PA+NPA group had similar pain relief. There was no significant difference between the NPA only group and No Analgesia group. The No Analgesia group had higher pain relief than the PA only group, however they started with a much lower pain score.

### Non-Pharmacological Pain Management Strategies

A total of 479 non-pharmacological interventions were received by 296 (45.8%) patients. The minimum number of non-pharmacological interventions received was one and the maximum was seven. Table 2 lists the different types of non-pharmacological analgesia by frequency of use, with heat application and distraction being the most frequently used. Encouragement to use non-pharmacological analgesia by nurses or doctors was reported as ‘often’ by 63 (21%) patients, ‘sometimes’ by 212 (72%) patients and ‘never’ by 20 (7%) patients.

**Table 2:** Frequency of use of different types of n*on-pharmacological analgesia in 296 adult ED patients with moderate to severe pain*.

| Type of non-pharmacological analgesia | Count | Percent |
| --- | --- | --- |
| Heat | 167 | 34.9% |
| Distraction | 107 | 22.3% |
| Cold pack | 62 | 12.9% |
| Walking | 17 | 3.5% |
| Meditation | 15 | 3.1% |
| Music | 13 | 2.7% |
| Deep breathing | 8 | 1.7% |
| Massage | 7 | 1.5% |
| Prayer | 4 | 0.8% |
| Relaxation | 4 | 0.8% |
| Imagery | 2 | 0.4% |
| Other | 73 | 15.2% |
| **Total** | **479** | **100.0%** |

### Factors associated with the use of non-pharmacological analgesia

Table 3 presents the significant univariable binary logistic models showing the association between the single variables with the receipt of non-pharmacological analgesia (non-significant variables are presented in supplementary table 2) and Table 4 presents the final multivariable logistic model.

**Table 3:** Significant single variable binary logistic models of the receipt of non-pharmacological analgesia.

| Variable | Sub-group | Unadjusted Odds ratio | 95% confidence interval of the odds ratio |  | Significance |
| --- | --- | --- | --- | --- | --- |
|  |  |  | Low | High |  |
| Gender | Female | Ref. |  |  |  |
|  | Male | 0.531 | 0.386 | 0.729 | <0.001 |
|  | Other | 0.899 | 0.125 | 6.456 | 0.916 |
| IRSAD |  | 0.997 | 0.994 | 0.999 | 0.040 |
| Age-adjusted CCI |  | 0.941 | 0.888 | 0.996 | 0.037 |
| Mode of arrival | Ambulance | Ref. |  |  |  |
|  | Walk in | 1.719 | 1.246 | 2.327 | <0.001 |
|  | Other | 3.299 | 0.259 | 36.866 | 0.332 |
| Streaming | Acute | Ref. |  |  |  |
|  | Resuscitation | 1.500 | 1.001 | 1.900 | P=<0.001 |
|  | Fast track | 3.654 | 0.069 | 15.987 | P=0.998 |
|  | Ramping | 9.635 | 6.958 | 15.888 | P=0.033 |
| Pre-hospital analgesia (Yes) |  | 0.517 | 0.365 | 0.732 | P=<0.001 |
| Daily average of access block time (per minute) |  | 0.996 | 0.994 | 0.997 | P=<0.001 |
| Daily Average of the Time to be Seen (per minute) |  | 1.008 | 1.001 | 1.015 | P=0.021 |
| Queensland Emergency Access Target (%) – Short Stay Ward |  | 0.987 | 0.978 | 0.995 | P=0.003 |
| Queensland Emergency Access Target (%) – Admission |  | 1.022 | 1.022 | 1.043 | P=0.031 |

CI = Confidence Interval, IRSAD = Index of Relative Socioeconomic Advantage and Disadvantage, CCI = Charlson Comorbidity Index, Ref = Reference Group, ED = Emergency Department. The Queensland Emergency Access Target is a measure of throughput, presented as a proportion of patients that are assessed, treated and discharged or admitted within four hours of arrival, with subgroups based on discharge location. Non-significant single variable binary logistic models are presented in supplementary table 2.

**Table 4:**
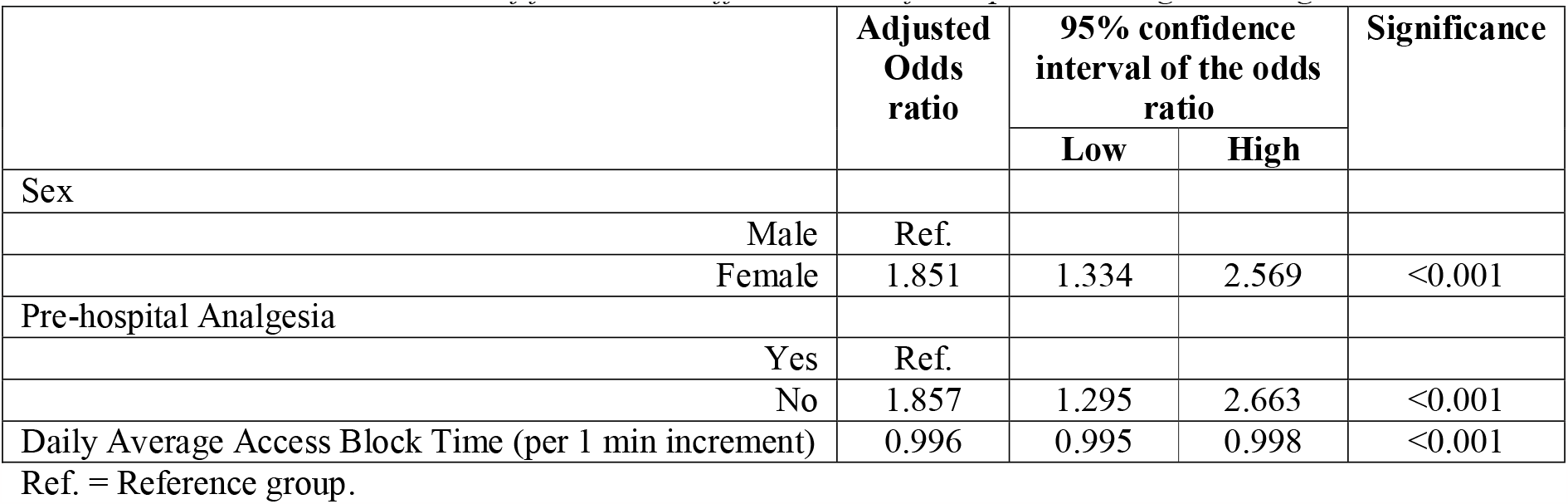
Final multivariate model of factors that affect the use of non-pharmacological analgesia in the ED.

|  | Adjusted Odds ratio | 95% confidence interval of the odds ratio |  | Significance |
| --- | --- | --- | --- | --- |
|  |  | Low | High |  |
| Sex |  |  |  |  |
| Male | Ref. |  |  |  |
| Female | 1.851 | 1.334 | 2.569 | <0.001 |
| Pre-hospital Analgesia |  |  |  |  |
| Yes | Ref. |  |  |  |
| No | 1.857 | 1.295 | 2.663 | <0.001 |
| Daily Average Access Block Time (per 1 min increment) | 0.996 | 0.995 | 0.998 | <0.001 |

Ref. = Reference group.

The use of non-pharmacological pain management in the ED is influenced by sex and the receipt of pre-hospital analgesia, with females and those who do not receive pre-hospital analgesia more likely to receive a non-pharmacological intervention. The workload of the hospital and its effects on the ED have a negative association with the use of non-pharmacological analgesia. These three variables combine to form a significant multivariable model accounting for 10.8% of the variation in the receipt of non-pharmacological analgesia. Age, mode of arrival, level and type of pain, and patient acuity did not significantly predict the use of non-pharmacological pain management in this sample.

### The association between reduction in pain and the use of non-pharmacological analgesia

Four factors were independent predictors of the amount of pain relief received in the ED with a linear regression (age-adjusted Charlson comorbidity score, the least pain experienced in the ED, the daily average of the time to be seen by a provider, and daily compliance with the four-hour rule) accounting for 62.5% of the variation of pain relief achieved. When the use of non-pharmacological analgesia was added to this linear model, it was significant (B0.331, 95%CI 0.032-0.063, p=0.030), however, accounted for only an additional 0.2% of the variation of the reduction in pain. (See Supplementary Table 3 for further detail).

## Discussion

The key finding in this study of adult ED patients with moderate to severe pain, is that nearly half (45.8%) of the patients received non-pharmacological pain management interventions as part of usual care. Where non-pharmacological interventions were received, they usually occurred in combination with pharmacological analgesia. Only a small proportion of patients (4%) received non-pharmacological pain management in isolation. In this study, the use of combination therapy was not more effective than pharmacological analgesia alone. However, the use of non-pharmacological analgesia was associated with a greater reduction in pain.

Non-pharmacological interventions are typically used for patients with lower pain scores and less severe injuries (18). The effectiveness of non-pharmacological interventions may be increased when multiple interventions are combined, particularly for elderly patients (19). In this cohort, a maximum of seven non-pharmacological interventions were reported by one patient. Additionally, combining non-pharmacological interventions with pharmacological analgesia can improve their effectiveness in managing pain for complicated conditions (20). This study demonstrated that the most common use of non-pharmacological analgesia is in combination with pharmacological analgesia and rarely on its own. As the population in this study all had moderate to severe pain on arrival (4/10 or greater) it would appear that, in these two EDs, non-pharmacological analgesia is used widely for all pain intensities.

Consistent with previous applications of SMT to the receipt of analgesia in the ED (11, 12), the use of non-pharmacological analgesia was influenced by multiple dimensions and domains of the model. While numerous differences in the population and conditions that used non-pharmacological analgesia were seen in bivariate analysis, the multivariable modelling showed that most of these differences were explained by only a few variables. In multivariable modelling, it was the sex of the patient (person dimension of the SMT), the receipt of pre-hospital analgesia (a component of symptom management strategies) and daily average access block time (environment dimension, an indirect measure of the functioning of the hospital influence over ED care) that influence the use of non-pharmacological analgesia. These factors are similar to those that influence the time to first analgesic medication in the ED (12).

Our findings that females utilise non-pharmacological pain management interventions more often than males echo those of previous studies in the ED and intensive care settings and during childbirth (21). Women are likely to combine non-pharmacological analgesia with low doses of standard pharmacological analgesia to avoid side effects. In the current study, the difference in prevalence of use of non-pharmacological analgesia between males and females was not explained by differences in pain severity as all participants had moderate to severe pain.

Our findings suggest that patients who have pre-hospital analgesia are less likely, than other patients with pain, to receive non-pharmacological analgesia in the ED. However, the reasons for this are unclear. In the setting of the current study, pre-hospital care providers have a wide range of pre-hospital analgesic options, including parental opiates and Ketamine. How these practices influence ED providers’ subsequent decisions about non-pharmacological analgesia are unknown, and warrant further research.

Hughes et al. have studied the ED environment and its influence on pain care extensively while defining this domain of SMT. Previous studies have shown that pain care suffers when the ED is busy, reflected in numerous metrics (11, 12, 22, 23). Again, in this work, we have shown that workload/occupancy of the hospital impacts pain care in the ED, with average access block time (a daily measure of the amount of time inpatients spend in the ED awaiting transfer to an appropriate inpatient care area) negatively affecting the provision of non-pharmacological analgesia.

Similar to the findings of this study, other studies reported using different types of non-pharmacological pain management interventions, such as applying heat, using cold packs, using acupuncture and acupressure, exercising muscles and joints, repositioning of body parts, restriction of movement, using massage, and applying hydrotherapy for chronic pain of musculoskeletal and chronic conditions (24, 25). The patients in this study used a wide variety of non-pharmacological analgesic modalities, many using more than one during their stay. The findings of this study would suggest that healthcare providers are integral to initiating non-pharmacological analgesia in the ED as has been seen in other settings (26, 27). However, further interventional studies should empower providers to utilise non-pharmacological analgesia, especially in conjunction with pharmacological analgesia.

## Limitations

The main limitation of this study was the use of a convenience sample rather than a random sample, which excluded patients who presented to the ED during late hours, overnight, and at weekends. In addition, only English-speaking patients were included, and patients with severe pain or cognitive impairments were excluded, which further limited the generalizability of the results.

## Conclusion

The use of non-pharmacological analgesia is common for patients presenting in moderate to severe pain to the adult ED. Doctors and nurses encourage the use of non-pharmacolgical analgesia, with over 90% of all patients using non-pharmacological analgesia encouraged to do so by their care providers. There are a wide variety of non-pharmacologcal analgesics used, most commonly in conjunction with pharmacological analgesia. There is a clear benefit in terms of pain reduction to the use of non-pharmacolgical analgesia and its use should be encouraged by care providers in the ED.

## Data Availability

Due to local regulations this data is not available

---

**Supplemental Table 1:** Additional demographic, presentation and clinical characteristics of 646 adult ED patients with moderate to severe pain, stratified by treatment group.

|  | Units | Total | Treatment Group |  |  |  | p |
| --- | --- | --- | --- | --- | --- | --- | --- |
|  |  |  | PA only | PA+NPA | NPA only | No Analgesia |  |
| Number of patients | n (%) | 646 | 322 (49.8) | 270 (41.8) | 26 (4.0) | 28 (4.3) |  |
| Private Health Insurance (Yes) | n (%) | 207 (32.0) | 221 (68.6) | 183 (68.0) | 18 (69.2) | 17 (60.7) | 0.858 |
| Ambulance | n (%) | 257 (39.8%) | 139 (43.2) | 86 (31.9) | 11 (42.3) | 21 (75.0) | <0.001 |
| Walk in |  | 386 (59.8%) | 182 (56.5) | 182 (67.4) | 15 (57.7) | 7 (25.0) |  |
| Other |  | 3 (0.5%) | 1 (0.3) | 2 (0.7) | 0 (0) | 0 (0) |  |
| ATS two | n (%) | 97 (15) | 54(16.8) | 35 (13.0) | 3 (11.5) | 5 (17.9) | 0.755 |
| ATS three |  | 369 (57.1) | 180 (55.9) | 156 (57.8) | 18 (69.2) | 15 (53.6) |  |
| ATS four |  | 165 (25.5) | 83 (25.8) | 70 (25.9) | 5 (19.2) | 7 (25.0) |  |
| ATS five |  | 15 (2.3) | 5 (1.6) | 9 (3.3) | 0 (0.0) | 1 (3.6) |  |
| Acute | n (%) | 416 (64.4) | 215 (66.8) | 160 (59.3) | 20 (76.9) | 21 (75.0) | 0.003 |
| Fast Track |  | 183 (28.3) | 77 (23.9) | 98 (36.3) | 4 (15.4) | 4 (14.3) |  |
| Resuscitation |  | 47 (7.3) | 30 (9.3) | 12 (4.4) | 2 (7.7) | 12 (4.4) |  |
| Admitted | n (%) | 428 (66.3) | 229 (71.1) | 170 (63.0) | 14 (53.8) | 15 (53.6) | 0.089 |
| Home |  | 206 (31.9) | 88 (27.3) | 93 (34.4) | 12 (46.2) | 13 (46.4) |  |
| Transferred |  | 12 (1.9) | 5 (1.6) | 7 (2.6) | 0 (0.0) | 0 (0.0) |  |
| Pre-hospital analgesia (Yes) | n (%) | 196 (30.3%) | 110 (34.2) | 62 (23.0) | 6 (23.1) | 18 (64.3%) | <0.001 |
| Daily average access block time | Mean minutes (SD) | 151.4 (110.8) | 171.5 (116.9) | 125.7 (94.6) | 134.6 (96.4) | 182.8 (142.4) | <0.001 |
| Daily average ED length of stay | Mean minutes (SD) | 270.7 (32.7) | 273.7 (32.5) | 267.5 (31.8) | 274.9 (27.3) | 263.5 (43.2) | 0.070 |
| Daily average of time to be seen | Mean minutes (SD) | 79.4 (23.6) | 78.1 (23.8) | 81.3 (23.3) | 86.1 (23.6) | 69.3 (20.5) | 0.018 |
| Queensland Emergency Access Target (%) – Short Stay | Mean (SD) | 50.2 (17.8) | 51.9 (16.8) | 47.8 (19.1) | 49.1 (14.8) | 54.4 (15.4) | 0.021 |
| Queensland Emergency Access Target (%) - Admission | Mean (SD) | 30.1 (7.9) | 29.3 (7.4) | 31.0 (8.3) | 28.9 (8.2) | 31.3 (8.0) | 0.042 |
| Queensland Emergency Access Target (%) - Discharge | Mean (SD) | 61.5 (11.3) | 62.0 (11.5) | 60.8 (11.3) | 58.2 (7.8) | 65.6 (9.8) | 0.061 |
| Queensland Emergency Access Target (%) – Total | Mean (SD) | 54.0 (8.2) | 54.2 (7.9) | 53.6 (8.7) | 52.1 (7.1) | 56.9 (7.0) | 0.128 |

ED = Emergency Department, PA only = patients who received pharmacological analgesia only, PA+NPA = patients who received both pharmacological and non-pharmacological analgesia, NPA only = patients who received non-pharmacological analgesia only, No Analgesia = patients who received neither pharmacological nor non-pharmacological analgesia. The Queensland Emergency Access Target is a measure of throughput, presented as a proportion of patients that are assessed, treated and discharged or admitted within four hours of arrival, with subgroups based on discharge location.

**Supplementary Table 2:**
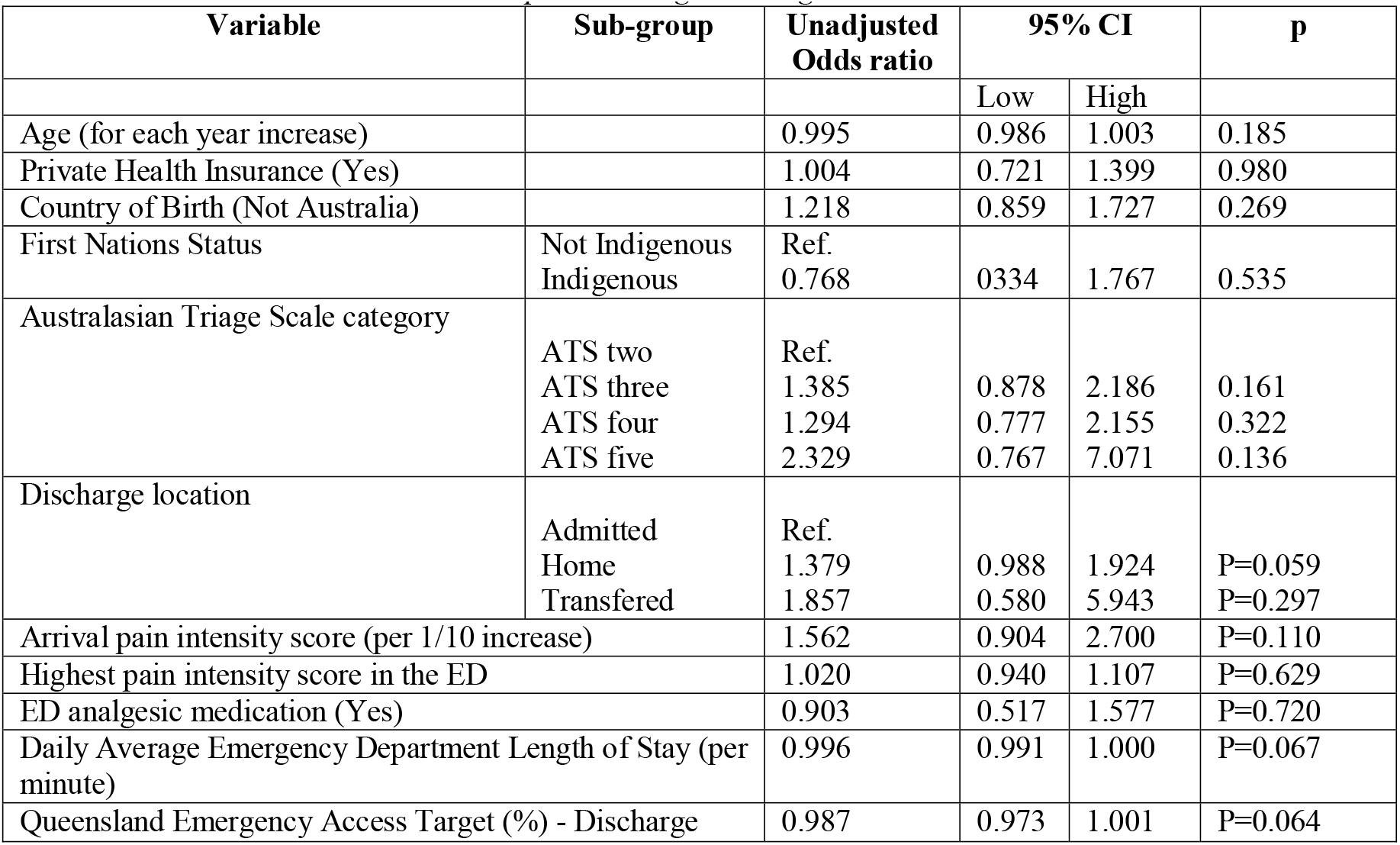
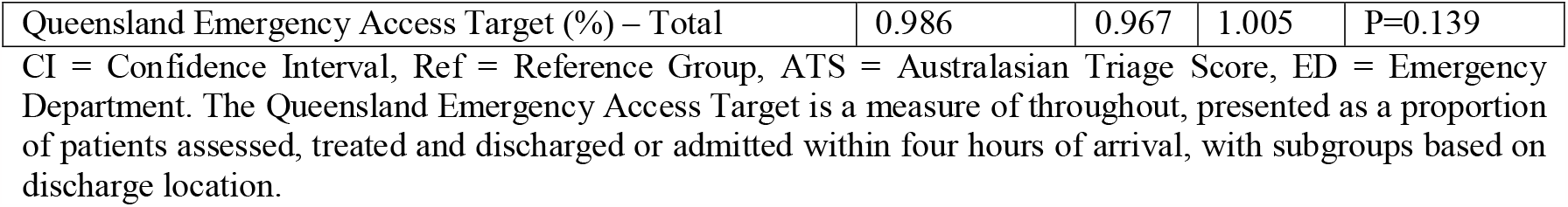
Non-significant single variable binary logistic models of the receipt of non-pharmacological analgesia.

**Supplementary Table 3:** Final multivariate model of factors that affect the reduction in pain experienced in the emergency department.

| | $\beta$ | 95% confidence interval of $\beta$ | | Significance |
| --- | --- | --- | --- | --- |
|  |  | Low | High |  |
| Age-adjusted Charlson Comorbidity Index | 0.062 | 0.008 | 0.115 | 0.024 |
| Least pain experienced | -1.050 | -1.116 | -0.984 | <0.001 |
| Daily time to be seen | -0.017 | -0.029 | -0.005 | 0.006 |
| Queensland Emergency Access Target (%) – Total | -0.058 | -0.092 | -0.025 | <0.001 |

